# Trans Cohorts Metabolomic Modulation Following Long-Term Successful Therapy in HIV-Infection

**DOI:** 10.1101/2021.07.14.21260539

**Authors:** Flora Mikaeloff, Sara Svensson-Akusjärvi, George Mondinde Ikomey, Shuba Krishnan, Maike Sperk, Soham Gupta, Gustavo Daniel Vega Magdaleno, Alejandra Escós, Emilia Lyonga, Marie Claire Okomo, Claude Tayou Tagne, Hemalatha Babu, Christian L Lorson, Ákos Végvári, Akhil C. Banerjea, Julianna Kele, Luke Elizabeth Hanna, Kamal Singh, João Pedro de Magalhães, Rui Benfeitas, Ujjwal Neogi

## Abstract

Despite successful combination antiretroviral therapy (cART), persistent low-grade immune activation together with inflammation and toxic antiretroviral drugs can lead to long-lasting metabolic adaptation in people living with HIV (PLWH). The successful short-term cART reported abnormalities in the metabolic reprogramming in PLWH, but the long-term consequences are unknown. This study investigated alterations in the plasma metabolic profiles by comparing PLWH and matched HIV-negative controls (HC) from Cameroon and India. We used untargeted and targeted LC-MS/MS-based metabolic profiling in PLWH with long-term (>5years) successful therapy in a trans cohorts’ approach. Advanced statistical and bioinformatics analyses showed altered amino acid metabolism, more specifically to glutaminolysis in PLWH with therapy than HIV-negative controls that can lead to excitotoxicity in both the cohorts. A significantly lower level of neurosteroids was observed in both cohorts and could potentiate neurological impairments in PLWH. The modulation of cellular glutaminolysis promoted increased cell death and latency reversal in pre-monocytic HIV-1 latent cell model U1, which may be essential for the clearance of the inducible reservoir in HIV-integrated cells. Our patient-based metabolomics and *in vitro* study, therefore, highlight the importance of altered glutaminolysis in PLWH that can be linked accelerated neurocognitive aging and metabolic reprogramming in latently infected cells.

## Introduction

Combination antiretroviral therapy (cART) can effectively block replication of human immunodeficiency virus type-1 (HIV). However, persistent low-grade immune activation together with inflammation, despite successful long-term treatment, and toxic antiretroviral drugs can lead to long lasting metabolic flexibility and adaptation in people living with HIV (PLWH) (1-3). Metabolic alterations have earlier been reported in HIV infection in treated and non-treated PLWH (1, 4-6). Furthermore, the metabolic adaptations associated with HIV infection are highly representative of immune dysregulation and inflammation related to accelerated aging (7, 8). However, there is a dearth of evidence about metabolic dysregulation due to long-term successful treatment in PLWH.

HIV regulates two essential immuno-metabolic pathways in the cell, namely glycolysis and glutaminolysis, to sustain the availability of biomolecules needed for viral replication in a cell-type dependent manner (9). The immune system is also adversely affected by HIV persistence and cell-to-cell spread that permits viral replication, despite on cART(10). This can further trigger transient or persistent metabolic changes that drive immune-senescence and accelerated aging in PLWH. Moreover, studies have shown altered glutaminolysis with high plasma glutamate levels in PLWH and that the modified glutaminolysis is responsible for the late immune recovery following cART (4).

In our recent untargeted metabolomics study on the COCOMO cohort from Denmark, we reported alterations in the amino acid (AA) metabolism as a central characteristic of PLWH with median of 13 years of therapy. This alteration was also more prominent in PLWH with metabolic syndrome (MetS) (3). However, reproducibility of metabolomic studies is challenging due to variability in metabolites linked with environmental factors and population-based heterogeneity in diet, gut microbiome, and lifestyle choices such as smoking or alcohol that have crucial impacts on individual metabolite composition and concentration (11-13). Finally, pre-analytical sampling errors, methodological errors, and informatics can influence the overall outcome and lead to cohort-specific effects.

This study investigates alterations in the plasma metabolic profiles by comparing PLWH on long-term cART and matched HIV-negative controls (HC) in two cohorts from low-and middle-income countries (LMIC), Cameroon and India, respectively, using untargeted metabolomics. We performed advanced statistical, bioinformatics and machine learning algorithms to identify the commonality between the cohorts associated with the cART in PLWH. We also used targeted metabolomics in a larger sample size of treatment naïve and experienced PLWH and HC from Cameroon and India to further validate our findings. Finally, to identify the metabolic state of the lymphocytic and pro-monocytic HIV-1 latent cell models, we performed *in vitro* studies by modulating the critical metabolic pathways identified in PLWH on cART in the presence and absence of the cART regimens. Our study provides a comprehensive metabolic profile of PLWH on cART, while HIV-latency in cellular models can shed light on the metabolic reprogramming in long-term successfully treated HIV infection and its potential role in accelerated aging in PLWH.

## Results

### Clinical Characteristics

We used a cohort of PLWH in Cameroon on cART (n=24, with a median duration of treatment of 11 years [IQR 8.00-13.25]) and HIV-negative controls (HC, n=24) (Table 1). All the patients were on a TDF/3TC/EFV-based regimen, non-smokers and omnivores. The only clinical parameter that achieved statistically significant difference (p = 0.007) between HC and cART patients was exercise.

**Table 1.** Clinical and demographic information.

|  | **Control** | **PLWH on cART** | **p-value** |
| --- | --- | --- | --- |
| **Number** | 24 | 24 |  |
| **Age in years, mean (95% CI)** | 48.00 (45.59-50.41) | 47.54 (42.54-52.5) | 0.86* |
| **Gender** |  |  | 1** |
| **Male, n (%)** | 12 (50) | 12 (50) |  |
| **Female, n (%)** | 12 (50) | 12 (50) |  |
| **CD4 current, median (IQR)** |  | 753.5 (540.5-827.8) | - |
| **CD8, median (IQR)** |  | 754.0 (483.2-955.0) | - |
| **Viral load, <40 copies/mL. n (%)** |  | 24 (100) | - |
| **Duration of the current regime in years, median (IQR)** |  | 11.00 (8.00-13-25) | - |
| **Alcohol consumption, n (%)** | 15 (62.50) | 17 (70.83) | 0.75** |
| **Exercise, n (%)** | 4 (16.67) | 14 (58.33) | 0.007** |
| **BMI, mean (95%)** | 27.88 (26.47-29.29) | 26.66 (23.90-29.41) | 0.42* |

\*Student t-test, \*\*Chi-square test,

### Impaired amino acid metabolism in PLWH on cART in the Cameroon Cohort

In plasma untargeted metabolomics, a total of 841 metabolites were detected, of which 46% (390/841) were lipids, 22% (188/841) were amino acids (AA) and 17% (143/841) were xenobiotics (Figure 1A). A low percentage of the metabolites were shown to be associated with environmental factors; diet, genetics, microbiome, lifestyle, and time of sampling as reported [11] (Figure S1). Out of all the detected metabolites, 122 metabolites differed significantly between PLWH on cART and HC (Mann-Whitney U test, p<0.05) of which 48% belonged to lipids (59/122) and 20% to AA (24/122). After correcting for multiple comparisons, 42 metabolites were statistically significant between PLWH on cART and HC (False discovery rate, FDR<0.1) (Figure 1A). Dimensionality reduction using these 42 metabolites showed an apparent clustering between HC and cART patients (Figure 1B). Among the 42 metabolites, 45% (19/42) were less abundant in PLWH on cART compared to HC. To identify mechanisms associated with HIV and cART, the differentially abundant metabolites having a Human Metabolome Database (HMDB) annotation (herein Mann-Whitney U test, p<0.05) were submitted to metabolite set enrichment analysis (MSEA) using Ingenuity Pathway Analysis (IPA). Based on the IPA (Z-score >2, FDR<0.05), the top identified pathways were concentration of lipids (n=15), synthesis of lipids (n=13), and production of reactive oxygen species (ROS) (n=12) (Figure 1C). The commonality of these pathways was the presence of glutamate while other AA such as arginine, cystine, and methionine were present in at least one pathway. These results showed a shift in lipid biosynthesis, immune activation of blood cells, and altered oxidative stress [production of reactive oxygen species (ROS) and hydrogen peroxide]. Metabolites within lipid metabolism exhibited the largest difference in PLWH on cART compared to HC (FDR<0.1) (Figure 1D). In this cluster, the changes appeared heterogeneous with a higher abundance of 7 metabolites and lower abundance of 12 metabolites in PLWH. Furthermore, AA, energy, and nucleotide metabolism were highly interlinked and upregulated in PLWH compared to HC. To complement the MSEA results, a balanced Random Forest (RF) algorithm was trained to predict HC and cART status of each metabolite based on Metabolon terms. Then, a permutation feature importance was applied to this machine learning technique. This analysis identified AA followed by lipids as the top pathways (Figure 1E) thereby confirming the importance of AA during cART in PLWH.

**Figure 1:**
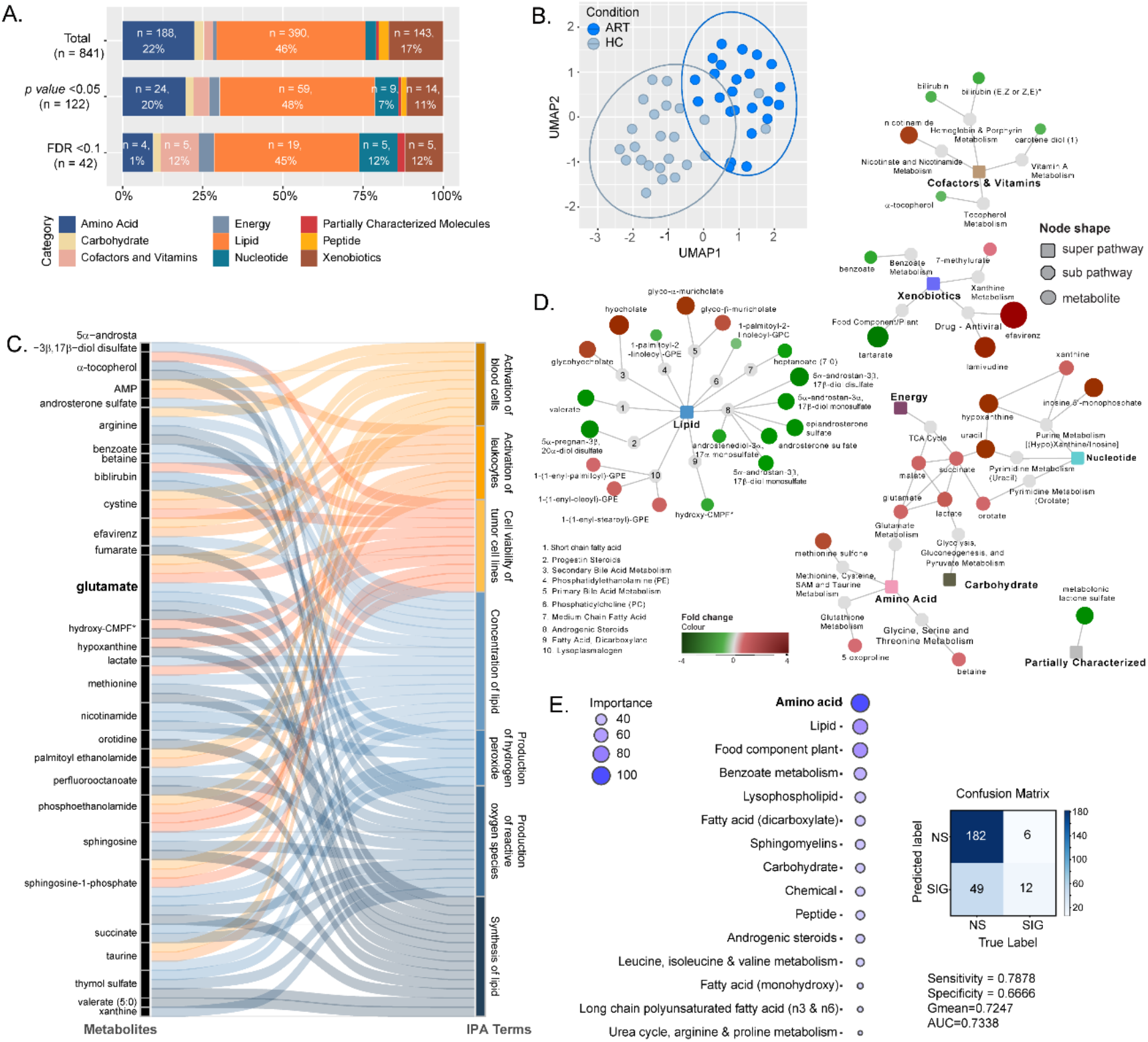
Untargeted metabolomics in the Cameroon Cohort highlight substantial metabolic alterations in cART compared with healthy controls. **(A)** Bar plots representing the proportion of super pathways and number of associated metabolites in total detected metabolites (n=841), metabolites with differential abundance between HC and PLWH on ART with p<0.02 (Mann-Whitney U Test, n=122) and FDR<0.1 (Mann-Whitney U Test, n=42). **(B)** UMAP visualization of 48 samples using metabolites differing between HC and PLWH on ART (Mann-Whitney U Test, FDR<0.1, n=42). Samples are colored by condition (light blue=HC; dark blue=cART). **(C)** Sankey Plot illustrating the most important contribution to the flow of glutamate associated pathways together with metabolites that are altered in cART patients. **(D)** Network of the metabolites significantly differing between HC and cART (Mann-Whitney U Test, FDR<0.1, n=42). Colored rectangular nodes represent super pathways, grey circles subpathways and colored circles single metabolites. The color gradient was applied depending on log2FC for each metabolite from green (decreased in cART) to red (increased in cART). The size of the bubble is proportional to log2FC. Edges connect each metabolite to its respective subpathway and each subpathway to their respective super pathway. **(E)** Bubble plot showing importance of Metabolon pathways in the prediction of metabolite association with cART status (left). Terms represented at the top of the figure are the most important for prediction. (RF, estimators: 500, class weight: balanced). Associated confusion matrix and classifier metrics are represented on the right.

### Altered neurosteroids as a common factor for the two cohorts from LMIC

To identify common biomarkers associated with HIV status and the impact of cART and strengthen our study, we combined the data from our Cameroon cohort with untargeted metabolomics analysis from an Indian cohort. The study design of the Indian cohort was similar using advanced bioinformatics and statistical analysis. Initially, we took the overlapping result of a linear classification model (PLS-DA), a machine learning model, RF and Mann-Whitney U test, performed in the two cohorts separately. The two separated RF models had a predictive accuracy of 97.9% in Cameroon and 100% in Indian cohort (Figure S2) after 10-fold cross-validation. We identified 14 (Mann-Whitney), 9 (PLS-DA) and 6 (RF) biomarkers differing between HC and ART in both Cameroon and Indian cohorts (Figure 2A). The overlap between all three methodologies identified six common metabolites to all methods (Figure S3). After removing the antiviral drug efavirenz, five metabolites were overlapping between the two cohorts: 5α−androstan−3α,17β−diol monosulfate, androsterone sulfate, epiandrosterone sulfate, metabolonic lactone sulfate, and methionine sulfone (Figure 2B). All the identified metabolites exhibited a similar trend between HC and cART in both cohorts thereby confirming their relevance. Furthermore, after correcting for confounders using multivariate linear regression, all metabolites were statistically significant (Figure S4, Table S4-S5). To further investigate the interactions of the metabolites related to HIV status, we performed a metabolite co-abundance network analysis in the Cameroon cohort based on significant positive pairwise correlation (Spearman, FDR<0.05) and used the Indian cohort for validation of the results. The network contained six communities found using Leiden algorithm (Figure 2C). The most central community (community 5, with the highest mean degree) showed 110 metabolites where the majority were lipids (85%), showing potential lipid dysregulation in cART (Figure 2D). Interestingly, four neurosteroids out of the five potential biomarkers; 5α−androstan−3α,17β−diol monosulfate, androsterone sulfate, epiandrosterone sulfate and metabolonic lactone sulfate, were in the same community (community 4) containing a total 123 metabolites. Metabolites from this community were less abundant in PLWH on cART compared to HC while community 6, containing glutamate, showed higher abundance. To validate the robustness of our complement biomarkers discovery approach, we used the biomarkers and their first neighbors from the co-expression network to separate HC and PLWH on cART in the Cameroon cohort. Based on hierarchical and consensus clustering, segregation of HC and PLWH on cART was observed in the Cameroon cohort (Figure 2E, left, Figure S5). As an independent validation, we performed a similar clustering in the Indian cohort (Figure 2E, right) using the same metabolite set and found an even better separation between HC and PLWH on cART than in the Cameroon cohort. Therefore, our data identified a set of correlated biomarkers, mainly neurosteroids, that were associated with PLWH on cART in two independent cohorts that could be linked to potential neurological impairments.

**Figure 2:**
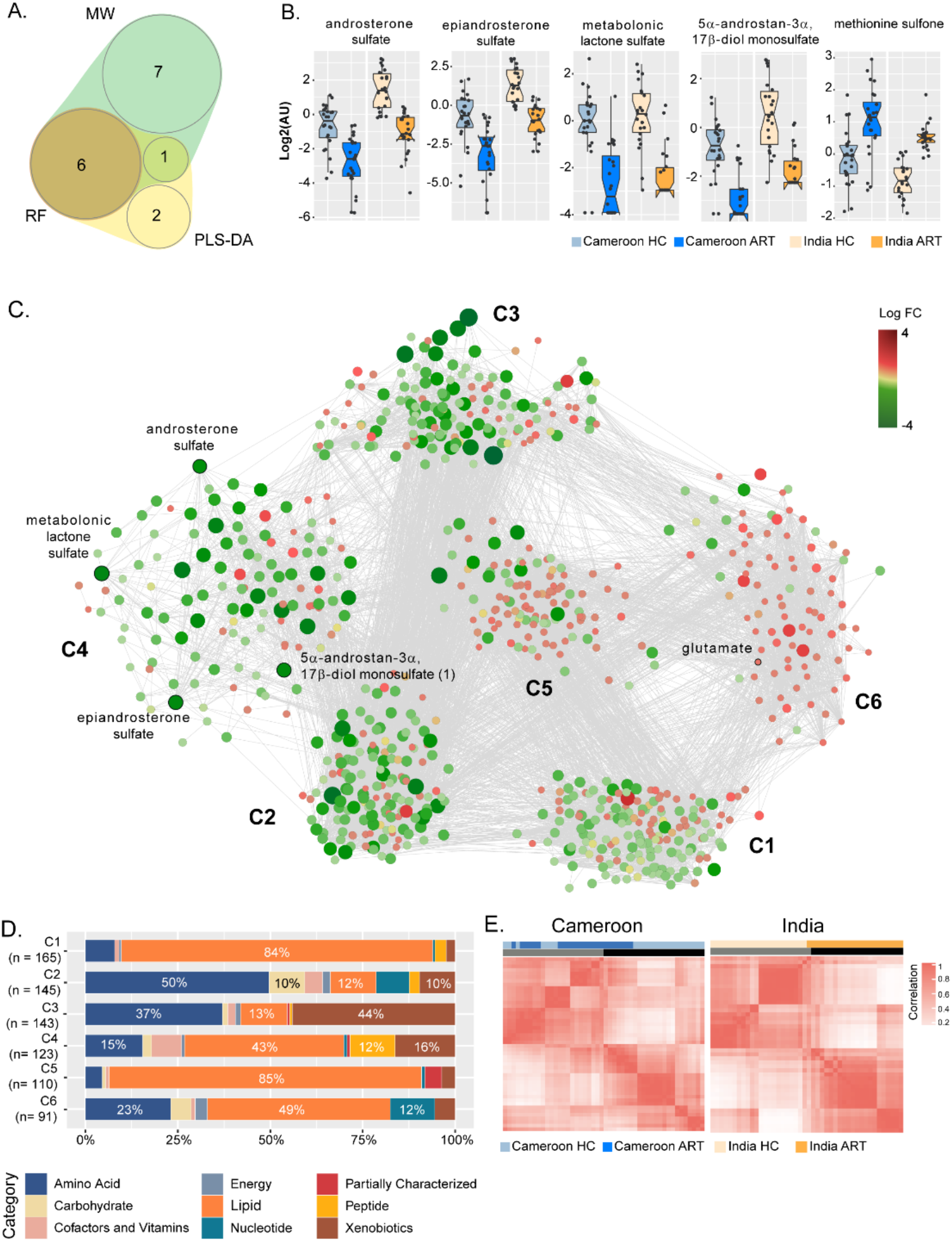
Identification of biomarkers associated with HIV-status and impact of cART compared to HC. **(A)** A 4-dimensional, quasi-proportional Venn diagram showing the number of overlapping metabolites (N = 6) differing HC/cART from three methodologies (Mann-Whitney U Test, RF and PLS-DA) in Indian and Cameroon cohort. Analysis was performed separately in Indian and Cameroon cohorts. **(B)** Boxplots of significant biomarkers shared by Indian and Cameroon patients: androsterone sulfate, epiandrosterone sulfate, metabolomic lactone sulfate, 5-α-androstan-3α,17β-diol monosulfate and methionine sulfone. In all the comparisons FDR<0.001 **(C)** Global association analysis network and identified communities. Potential biomarkers and glutamate are indicated. **(D)** Bar plots representing proportion of super pathways and number of associated metabolites in communities (n_c1=165, n_c2=145, n_c3=143, n_c4=123, n_c5=110, n_c6=91). **(E)** Consensus matrices of potential biomarkers and first neighbors in HC and cART. Data were log-transformed and z-score transformed.

### Targeted metabolomics in a larger cohort identified altered amino acid metabolism in two independent cohorts with differential mechanisms

To validate the importance of AA (as observed in Figure 1), we performed targeted metabolomics for AA in 90 PLWH on cART, 78 HC, and 45 untreated HIV-infected patients with viremia from Cameroon (n=123) and India (n=90). Even though the samples were run together, there was a clear cohort effect (Figure S6). Therefore, the subsequent analyses were performed on the cohorts separately. Eight AA were altered in the Cameroon cohort and 13 AA were altered in the Indian cohort between PLWH on cART and HC. Among these, 6 AA were overlapping between both the cohorts (Figure 3A, Figure S7), of which five essential AA including methionine, phenylalanine, threonine, valine, and tryptophan were significantly lower (FDR < 0.1) in PLWH on cART (Figure 3B-F). Only glutamate was significantly higher in PLWH on cART compared to HC in both cohorts. Interestingly, glutamate was significantly higher in untreated HIV-infected PLWH than HC in the Indian cohort but not in the Cameroon cohort (Figure 3G). Taken together these results indicate the importance of glutamate and AA in PLWH on cART. To assess the size of the difference over significance, the effect size calculation was performed based on sample mean difference (ART-HC) using Glass delta (D) (Table S6). Overall, similar direction of the effect was observed in both cohorts (Figure 3H-I). Glutamate tests were medium (D = 0.62) and small (D = 0.25) in Cameroon and Indian cohort, respectively, and showed a greater mean in ART compared to HC. The largest effect sizes were observed in the Indian cohort on tryptophan (D = -1.07), serine (D = -0.84) and methionine (D = -0.81) (Figure 3I). Overall, these results indicate alterations in glutaminolysis as a common phenomenon in long-term treated individuals and that glutamate, glutamine and GABA plays an important role in metabolic reprogramming.

**Figure 3:**
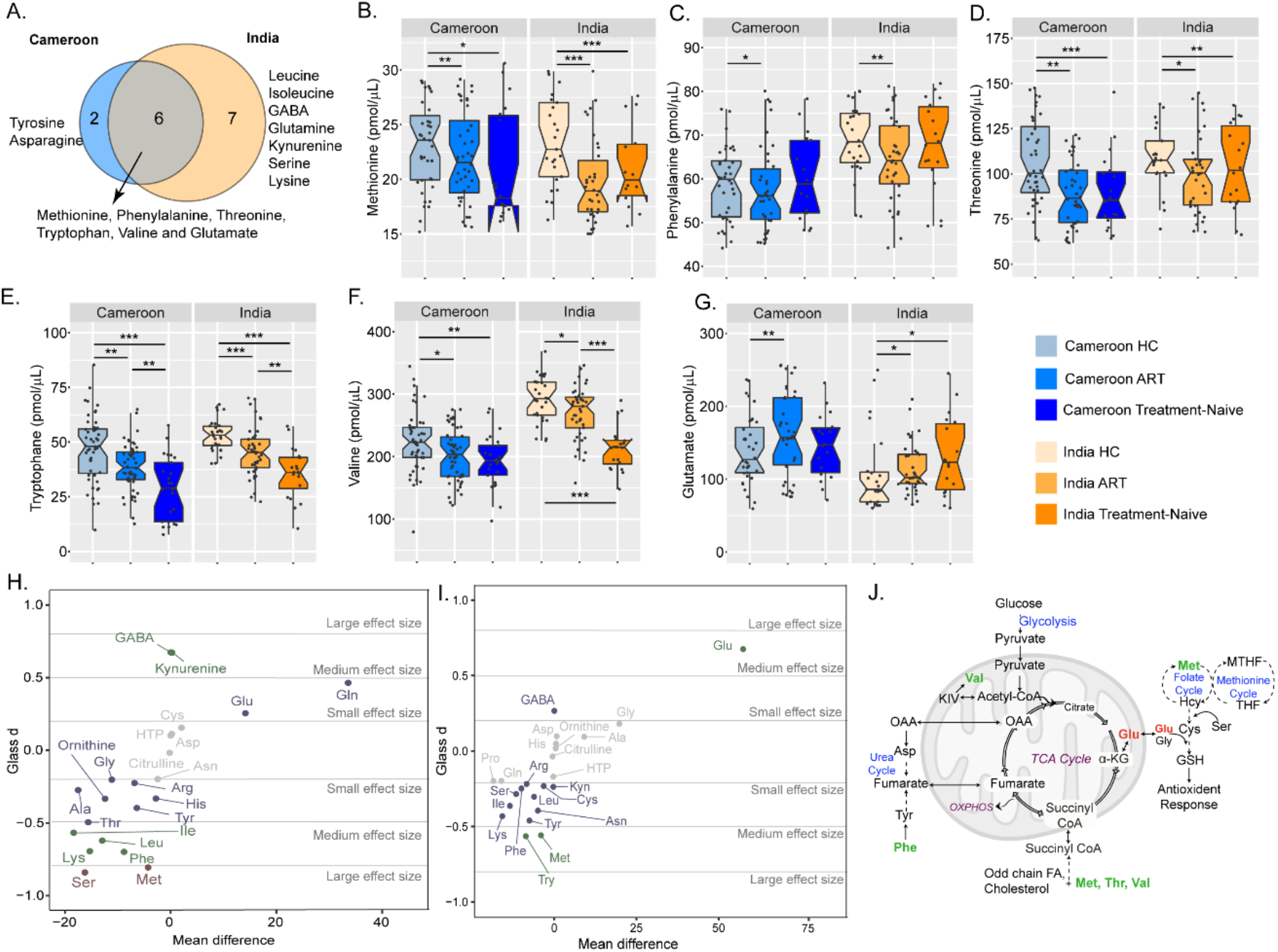
Targeted amino acid in the larger HIV-1 cohorts from Cameroon and India. **(A)** Venn diagram representing the overlap of AA significantly differing in HC compared to cART between Cameroon and Indian cohort. **(B-G)** Box plots showing the abundance of six significant AA between HC and cART (Mann-Whitney U test, FDR<0.1) (* FDR<0.1, ** FDR<0.05, *** FDR < 0.01): methionine **(B)**, phenylalanine **(C)**, threonine **(D)**, tryptophan **(E)**, valine **(F)**, and glutamate **(G)** in HC, and PLWH on cART and treatment-naive patients from Cameroon and Indian cohorts. **(H and I)** Scatter plot of AA mean difference by effect size (Glass d) in Indian **(H)** and Cameroon **(I)** cohorts. Dots are colored based on effect size (red = large, green = medium, blue = small). **(J)** Schematic representation of the altered AA linked with the key metabolic pathways.

### Role of altered glycolysis and glutaminolysis in HIV-latency cell models

We observed alterations in the AA metabolism in PLWH on cART. Therefore, we aimed to study the effect of glycolysis and glutaminolysis during HIV latency, both of which are important for supplement and utilization of AA and energy production by the TCA cycle (Figure 3E). During cART, HIV persists in latent reservoirs both from monocytic and lymphocytic lineages. Earlier studies have shown how metabolic regulation may be a key factor regulating HIV infection where alterations in glycolysis have a large effect (14, 15). To characterize alterations in cellular metabolism during the steady-state of latent HIV infection, we performed quantitative proteomic analysis using LC-MS/MS in pro-monocytic latent cell model U1, and lymphocytic latent cell model J-lat10.6 together with their respective uninfected parental cell lines, U937 and Jurkat. The steady-state modifications in the cell lines upon entering HIV latency showed significant alterations in the biosynthesis of AA and TCA cycle (Figure 4A). First, we used 6-diazo-5-oxo-L-norleucine (DON) and 2-deoxy-D-glucose (2-DG) to block glutaminolysis and glycolysis respectively, followed by a latency reversal agent prostratin to understand the roles of these pathways during latency reversal in J-Lat10.6 and U1 cell lines. Both latency cell models, J-Lat10.6 and U1 exhibited a decrease in viability when treated with 2-DG or DON compared to Jurkat and U937 cells (Figure 4B). As expected, prostratin, a protein kinase C agonist, activated the latent reservoir in both cell lines (Figure 4C, 4D). Interestingly, blocking glutaminolysis using DON alone was able to activate the latent reservoir in U1 cells (Figure 4D) but not in J-Lat10.6 cells (Figure 4C). To further explore the alterations concomitant with blocking glutaminolysis, we performed quantitative proteomic analysis using LC-MS/MS in U1 and U937 cells following treatment with DON, prostratin, and DON+prostratin. Because differences in the steady-state protein levels are observed between the two cell lines, we corrected the effect of U937 in U1 cells and performed differential abundance analysis and protein set enrichment analysis (Figure 4E). Addition of prostratin alone in U1 cells showed alterations in KEGG metabolism pathways such as carbon metabolism, TCA cycle and AA metabolism (p<0.1, Figure S8), which could be linked to latent HIV reservoir activation. Exposure to DON showed dysregulation of several metabolic pathways including glycolysis/gluconeogenesis, TCA cycle, sulfur metabolism, valine, leucine, isoleucine degradation and oxidative phosphorylation (OXPHOS). Several proteins of OXPHOS complexes I, III and IV, displayed lower abundance in U1+DON compared to U1 control cells (Figure 4F). To confirm this, we performed western blot using human OXPHOS antibody cocktail that detects the components of complex-V (ATP5a), complex-III (UQCRC2), complex-II (SDHB), complex-IV (COX II) and complex-I (NDUFB8). Since we could not differentiate complex -I and -IV in all our western blots, we considered them together in our analysis. While we did not observe any statistically significant differences in the protein levels of the OXPHOS complexes, visible decrease in expression of complexes -III, -II, -I and -IV but not complex-V were observed upon DON treatment both in presence and absence of prostratin (Figure 4H). Overall, our experimental data correlated with the proteomics data and is suggestive of suppression of OXPHOS upon blocking glutaminolysis in HIV latently infected promonocytic cells. Since changes in OXPHOS are often linked to unbalanced redox homeostasis, we measured the total cellular ROS levels in U1 cells following treatment with prostratin, DON and DON+prostratin. We observed a decrease in cellular ROS levels in the presence of prostratin compared to the control. However, inhibition of glutaminolysis by DON did not alter ROS levels compared to control cells (p>0.05, ns). Conclusively, it can be postulated that inhibition of glutaminolysis can compensate for the effect of HIV latency by altering oxidative stress. Therefore, ROS may not be the obligate driver of the metabolic reprogramming in the cell models of HIV-latency.

**Figure 4:**
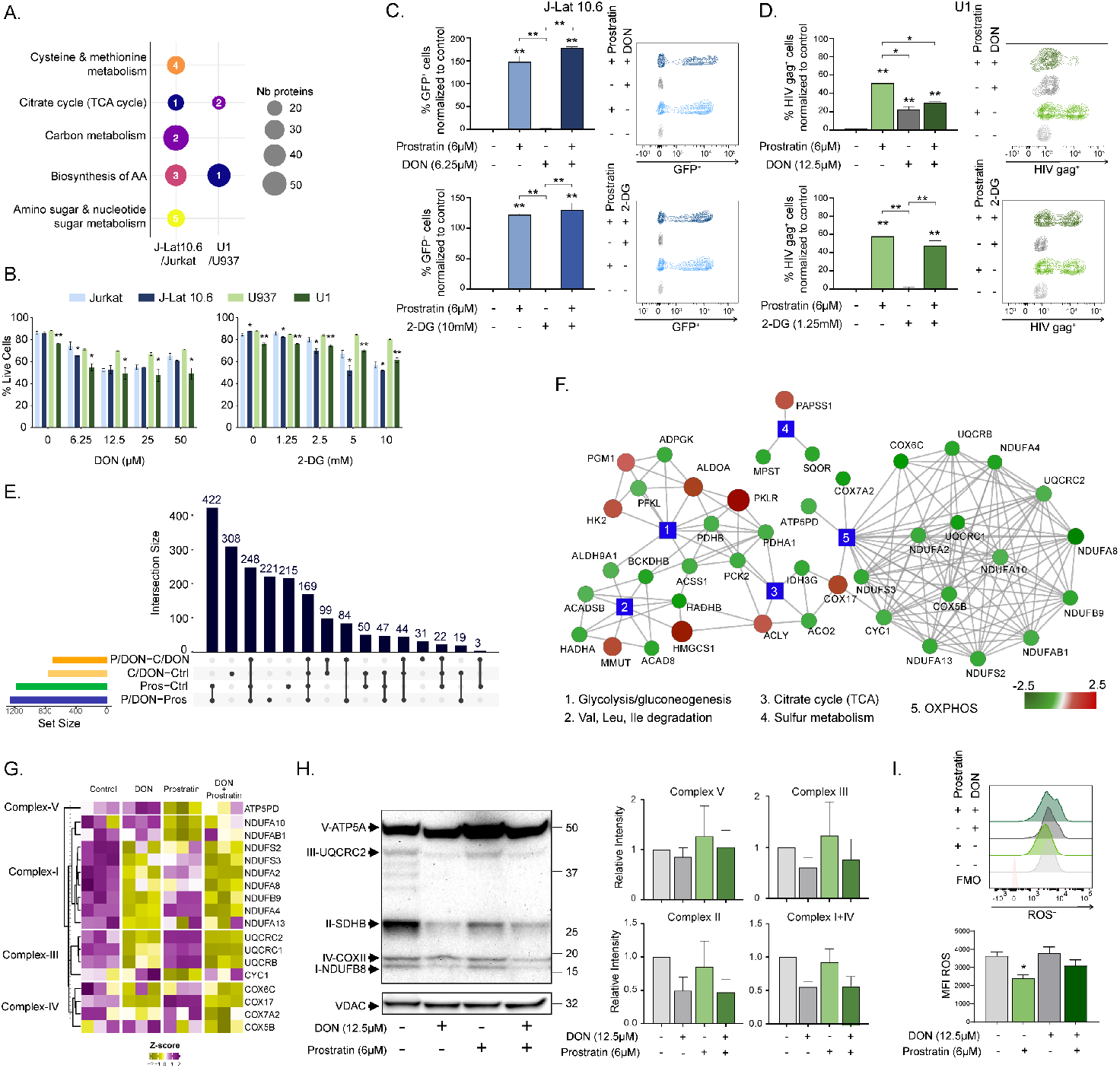
Effect on cellular metabolism in latency cell models. **(A)** Steady-state metabolic alterations in the HIV-latency cell models, J-Lat10.6 and U1, compared to Jurkat and U937, respectively. MSEA using the KEGG Metabolism with FDR<0.05 shown as bubble plots. Size of the bubble represents the number of proteins and the number, the rank of the pathway based on FDR. **(B)** Viability of latency cell models J-Lat10.6 and U1 compared to respective parental cell lines Jurkat and U937 during 48h DON or 2-DG treatment measured using flow cytometry. **(C, D)** HIV latency activation using prostratin (6μM) together with DON (6.25μM) or 2-DG (10mM) in J-Lat10.6 cell line **(C)** and DON (12.5μM) or 2-DG (1.25mM) in U1 cell line **(D). (E)** Upset plot of proteins with differential abundance between control vs. DON (C/DON-Ctrl), control vs. prostratin (Pros-Ctrl), prostratin vs. DON+prostratin (P/DON-Pros) and DON+prostratin vs. DON (P/DON-C/DON) in U1 cells corrected for U937 cells. **(F)** Network of the proteins differing significantly between U1 and DON treated U1 (FDR<0.1, n=758). Color gradient was applied on log2FC from green (decreased in U1-DON) to red (increased). **(G)** Heatmap showing OXPHOS proteins levels in U1 treated with DON and prostratin (n=18). Proteins were selected based on comparison U1 vs U1-DON (LIMMA, FDR<0.1) and their associated with OXPHOS KEGG pathway. Proteins were separated based on their complexes (from I to V). **(H)** Western blot analysis of OXPHOS proteins during DON treatment in U1 cells (left). Representative blot of three independent experiments is shown here. Quantification of western blot represented as bar graphs represented with mean±SEM (right). **(I)** Measurement of ROS in latency cell model U1 during DON treatment. Statistical analysis was performed using unpaired t test or Mann-Whitney U test (* p<0.05, and ** p<0.001) and represented with mean±SEM. All experiments were performed in three independent replicates.

### 2-DG and DON modulates intracellular metabolite levels independent of ART regimens in U1 cells

Thereafter, we sought to evaluate the effect of 2-DG and DON during latency reversal in the presence of cART regimens; tenofovir disoproxil fumarate (TDF) + lamivudine (3TC) + efavirenz (EFV) (TDF+3TC+EFV), and zidovudine (AZT) + 3TC + EFV (AZT+3TC+EFV) prevalently used in low-income countries. Cellular cytotoxicity of cART regimen was evaluated using Alamar blue assay (Figure S8). In our promonocytic latency model, U1, prostratin increased HIV gag protein expression independent of the cART regimens. However, both 2-DG and DON in combination with prostratin reduced the expression of gag (Figure 5A). The increased gag expression by DON alone, as observed earlier, was detected independently of cART. Furthermore, to understand how 2-DG and DON modulate intracellular metabolite levels we used metabolite measurement kits to measure glucose, lactate, and glutamate levels during latency reversal under cART pressure in U1 cells (Figure 5B). Similar trends could be seen for the measured metabolites both in the presence and absence of cART (Figure 5C). Intracellular glucose levels decreased during prostratin treatment while 2-DG and DON increased glucose levels both with and without prostratin compared to control. Intracellular lactate levels were reduced during prostratin treatment both with and without the inhibitors compared to control. Interestingly, when treating U1 with prostratin and DON, lactate levels significantly increased compared to DON alone, independent of cART. Furthermore, prostratin increased intracellular glutamate levels both alone and with 2-DG or DON in absence of cART while under cART pressure the increase was only seen using prostratin or 2-DG alone. As 2-DG reduced glutamate levels, this increase could be caused by restitution of glutamate levels by prostratin.

**Figure 5.**
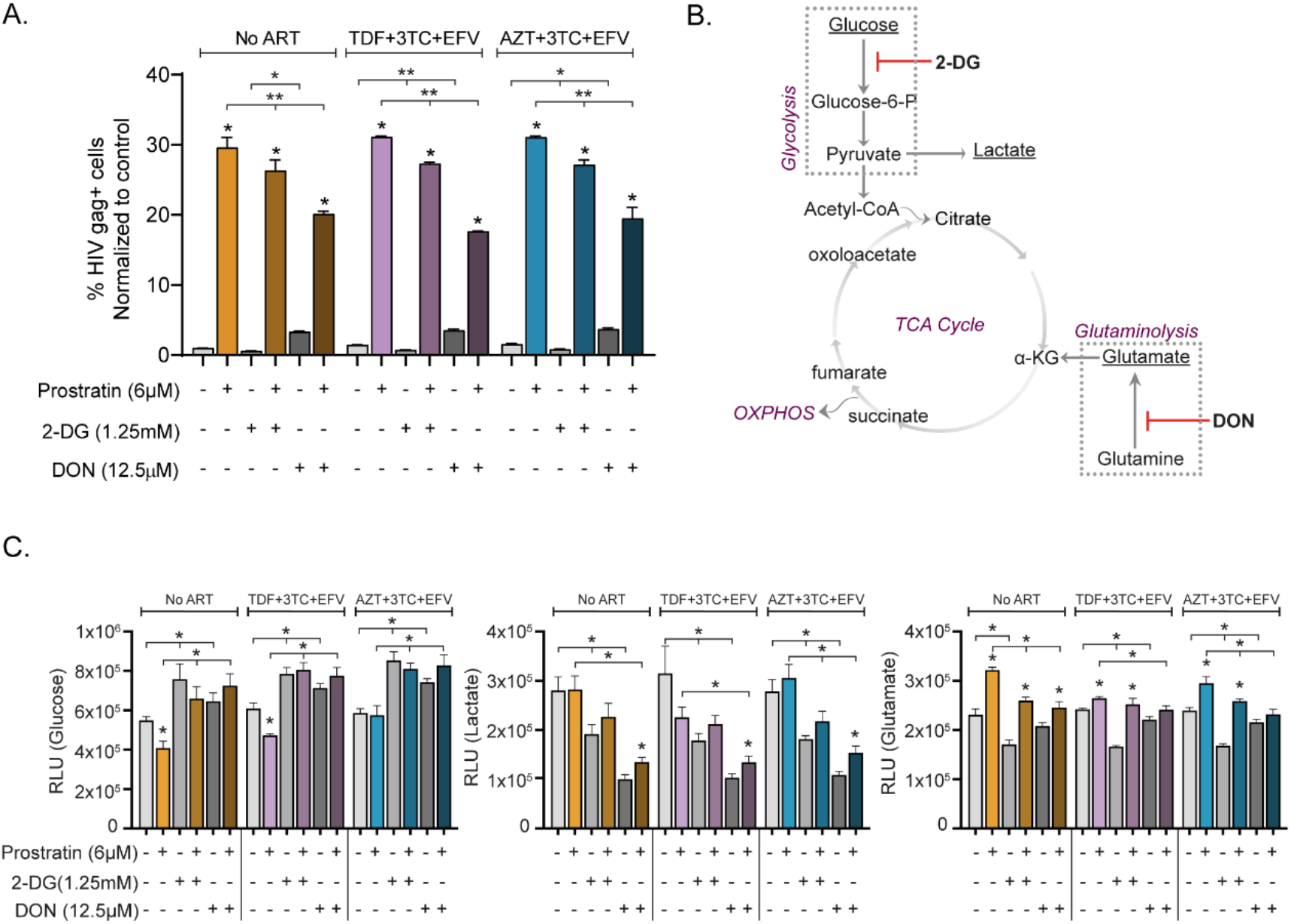
Effect of cART regimens on HIV activation during inhibition of glutaminolysis: Effect of cART regimens (TDF+3TC+EFV and AZT+3TC+EFV) in combination with 2-DG and DON on HIV latency activation in U1 cells. **(A)** Production of HIV gag during cART treatment in the presence of metabolic blockers 2-DG or DON. **(B)** Schematic showing the effect of 2-DG and DON on metabolic processes. **(C)** Effect of cART regimens on intracellular glucose, lactate, and glutamate levels when treated with 2-DG or DON during latency activation. All experiments were performed in three independent replicates. Statistical analysis was performed using Mann-Whitney U test (* p<0.05, and ** p<0.001) and represented as mean±SEM.

## Discussion

Our study identified altered plasma AA profiles in two HIV cohorts of PLWH, from Cameroon and India, on successful cART. The common factor in this trans-cohort study was lower level of essential AAs methionine, phenylalanine, threonine, valine, and tryptophan and elevated glutamate in PLWH on cART compared to HC. Significantly lower levels of neuro-steroids like 5α−androstan−3α,17β−diol monosulfate, androsterone sulfate were observed in both cohorts. Modulation of cellular glutaminolysis increased cell death and latency reversal in promonocytic HIV latency cell model U1. Therefore, our study highlights the importance of altered glutaminolysis in PLWH that can be linked to accelerated neurocognitive aging and metabolic reprogramming in latently infected cells.

The metabolic network of AA is co-regulated and highly complex. Therefore, we conducted a trans-cohort metabolic profile analysis using cutting-edgestatistical analysis, machine learning algorithms, together with network analysis to identify metabolic features in PLWH. Our findings indicated an in-depth dysregulation of AA metabolism in long-term treated PLWH. Earlier studies on untreated and short-term treated PLWH (up to 36 months) on smaller cohorts from USA, Netherlands, and Spain have shown disruption of AA metabolism during infection (16-18). In our study, we showed that AA dysregulation is a persistent feature even with more than five years of treatment. This result is in line with our recent large-scale metabolomics study from the Copenhagen Comorbidity in HIV-infection (COCOMO) cohort with a median duration of 13 years treatment. In the COCOMO cohort we observed altered AA metabolism in PLWH both with and without metabolic syndrome (MetS) compared to the HC. Alterations in AA such as tryptophan, glutamine, glutamate, phenylalanine, arginine, aspartate, and threonine have been closely linked to HIV and cART-induced metabolic complications as well as oxidative stress (19, 20).

In our current study, we observed increased plasma levels of glutamate in PLWH on cART compared to HC in both cohorts while it was only in the Indian cohort that treatment naive PLWH had increased glutamate levels. Interestingly, in the Cameroon cohort treatment naive PLWH had median glutamate levels like HC. One of the potential reasons could be that pre-therapy PLWH from India had a much lower CD4 count [median (IQR): 300.5 (213.2-527.0) cells/uL] compared to Cameroon cohort [median (IQR): 495.0 (382.0-558.0) cells/uL]. Possibly a severe depletion of CD4^+^ T-cells can influence the metabolic environment and cause detrimental effects. This hypothesis is further supported by our earlier studies showing PLWH with MetS have significantly lower nadir CD4^+^ T-cell count (3).

Glutamate displays remarkable metabolic versatility in several metabolic pathways including both AA synthesis as well as degradation. However, glutamate-mediated excitotoxicity is the primary contributor to age-related neurodegenerative disorders (21). Elevated plasma glutamate in PLWH may cause loss of lymphocyte and macrophage function (22). The primary source of extracellular glutamate in HIV infection includes release of intercellular glutamate due to cell death, macrophages/microglia activation, and disrupted neurotransmitter clearance (23). Metabolic tracer experiments demonstrated considerable changes in the glutamine metabolism with increased secretion of glutamine-derived glutamate from HIV-infected CD4^+^ T-cells (24). An earlier study also reported that HIV infection enhances glutamate production in human monocyte-derived macrophages that may be an important link to HIV-associated dementia (25). In pre-clinical studies of EcoHIV Murine Model of HIV-associated neurocognitive disorders (HAND), the inhibition of the glutaminase (GLS) with DON or a DON prodrug JHU083 reversed the impaired cognitive function, indicating the role of the glutaminolysis in HAND (26, 27). In our study, we observed increased plasma glutamate in both the cohorts and low neuroactive steroids in PLWH with therapy compared to the HC. A recent study in PLWH with high and low depressive symptoms also reported a reduced level of neuroactive steroids in participants with high depressive symptoms (28) indicating that depression severity associate with lower levels of neuroactive steroids. Interestingly, neuroactive steroids have been shown to regulate glutamatergic neurotransmission (can bind to receptors for glutamate) as well as behavioral actions (29). Taken together, it can be hypothesized that increased plasma glutamate and decreased neurosteroids in PLWH following successful therapy have the potential to develop neurocognitive impairment and depressive disorders that may need the clinical intervention.

Metabolic reprogramming occurs to ensure energy availability and to elicit an appropriate immune response upon pathogen encounter. Susceptibility to HIV is partially regulated by activation stage and the metabolic activity of a cell where elevated OXPHOS and glycolysis favors infection in lymphocytes (30, 31). Even as the main HIV reservoir is believed to reside in long-lived lymphocytic cells, latently infected monocytes and macrophages can persist over time and facilitate spread during cART. In this study, we show the effect of inhibition of glutaminolysis on latent reservoir in monocytic cells. On a metabolic level, this results in a reduction of proteins involved in OXPHOS while increasing proteins involved in glycolysis, proteinogenic branched-chain AA degradation (valine, leucine, and isoleucine) and TCA cycle. Previous studies have shown increased glutamine metabolism in latently infected cells (32-34). Specifically, latently infected macrophages use glutaminolysis as a primary energy source in addition to fatty acid and glucose used by their uninfected counterparts (35). Furthermore, macrophages carrying latent HIV have a compromised TCA cycle that induces lipid accumulation and OXPHOS and enlarged mitochondrion (35). In our study, glutaminolysis inhibition is accompanied by altered metabolite levels with increased intracellular glucose and reduced glutamate and lactate levels irrespective of cART treatment. Uptake of glucose through Glut1 regulates susceptibility of HIV-1 and lymphocytes carrying latent HIV-1 have also been proposed to express OX40 together with Glut1 (36, 37). Elevated glycolytic activity, Glut1 expression and immune activation are also seen in people living with HIV and factors needed for virion production (38, 39). Herein, we did not see any effect on latency reversal when inhibiting glycolysis. Therefore, while glycolysis is an important factor for HIV-1 entry and replication, latency reversal is somewhat dependent on glutaminolysis. Furthermore, proteins involved in mitochondrial respiration were more abundant in latent monocytic U1 cells than the parental U937 cell line indicating that OXPHOS proteins may play a role in HIV persistence. Therefore, a unique metabolic environment may be induced by the virus to maintain a transcriptionally inactive state. By inhibiting glutaminolysis, cellular metabolism can be altered and thereby force transcriptional activation of latent HIV-1.

In conclusion, our present study based on cohorts (India and Cameroon) indicated altered AA metabolism and more potentially a switch in glutaminolysis as the alternative pathway for energy production following a long-term antiretroviral therapy, corroborating our earlier study (3). Altered glutaminolysis with long-term treatment and its association with metabolic syndrome (3), diminished immune recovery (4), and glutamate excitotoxicity mediated neuro-cognitive impairments can lead to increased co-morbidities and accelerated aging in PLWH with successful therapy.In addition, a decrease in neurosteroids causes major depressive syndrome (28) leading to diminished quality of life despite successful treatment. Our study also provided evidence displaying the cross-talk between glutaminolysis, TCA cycle and OXPHOS in HIV-latent cell model being more specific to pro-monocytic U1 cells that potentially is linked with apoptosis as well as latency reversal. Increased knowledge about the co-regulation of interconnected metabolic pathways in the context of HIV infection and therapy can provide new targets for future therapeutic interventions both for improving metabolic health as well as other metabolic disorders in PLWH. It can also reveal a potential for clearing the latent reservoir by modulating the cellular metabolic pathways as a novel strategy for functional cure.

## Methods

### Study population

This study included three groups of individuals in Cameroon: HIV-1 infected individuals on combination ART (cART, n=50), untreated HIV-1 infected individuals with viremia (treatment-naive, n=25) and HIV-negative individuals (HC, n=50). The study groups are age-and BMI-matched with comparable gender proportions between males and females. Whole blood and plasma were collected from Yaounde University Teaching Hospital, Cameroon. Additionally, participants from an Indian cohort were included PLWH on cART (n=41), treatment naïve (n=20), and HC (n=30), as reported earlier (2).

### Cell lines

For this study latency cell models J-Lat10.6 (AIDS reagent program) and U1 (AIDS reagent program) were used together with their respective parental cell lines Jurkat (AIDS reagent program) and U937 (Kindly provided by Helena Jernberg Wiklund, Uppsala University).

### Untargeted and targeted metabolomics

Plasma untargeted metabolomics was performed at Metabolon, Inc. (North Carolina, USA) on a selection of individuals from the Cameroon cohort including HC (n=24) and cART (n=24), as previously described (1, 40). In a larger cohort targeted metabolomics for amino acids was performed using LC-MS/MS method with reference amino acids as control at the Swedish Metabolomics Centre (Umeå, Sweden). For detailed method and quality control please see supplementary method. Untargeted proteomics was performed at the Proteomics Biomedicum facility, Karolinska Institutet as described by us recently (41).

## Bioinformatics and statistical analysis

### Statistics

The Non-parametric Mann-Whitney U, Spearman correlations as well as linear regression were performed using R (https://www.r-project.org/). Machine learning models were built using R packages Boruta for feature selection (42), randomForest with 10-fold cross validation, and python scikit-lean (43) and verified using confusion matrices and the Area under the Receiver Operator Curve (ROC). PLD-DA plot was made using ropls (44). For proteomics, preprocessing was performed as described previously (41) and differential abundance analysis using R package LIMMA (45) using following design : [∼ cell*p + cell*don + cell:p:don]. False discovery rate (FDR) was applied for multiple comparisons correction. Effect sizes were calculated using the d_glass function from the R package effectsize to compensate for relevant differences in the standard deviations.

### Pathways and clustering analysis

Pathway Analysis software (IPA) software (QIAGEN Inc., https://www.qiagenbioinformatics.com/products/ingenuitypathway-analysis) and Metabolon terms were used in metabolomics data for pathway analysis. For proteomics, curated Metabolic KEGG Human 2019 libraries and enrichr function from gseapy python package were applied (version 0.10.3; https://github.com/zqfang/gseapy). Cutoff for pathways were set to FDR<0,05. For association analysis, significant positive correlations network (Spearman, FDR < 0.05) was built using python igraph (https://igraph.org/python/). Community analysis was done using Leiden algorithm and most central community found using mean degree (46). Complete methodology was described before (41). Cluster analysis was conducted using R package ConsensusClusterPlus with following metrics [algo : hierarchical clustering, distance : spearman, pitem : 0.8, reps : 1000](47).

### Visualization

Dimensionality reduction was performed using UMAP (48). R package ggplot2 (49), ggalluvial (50), nVennR (51), VennDiagram (52), UpsetR (53) and Complexheatmap (54) were used to generate figures. Cytoscape ver 3.6.1 was used for network representation (55).

### Laboratory Experiments

#### Cytotoxicity assays

Toxicity of cART regimens (TDF+3TC+EFV and AZT+3TC+EFV) or 2-DG and DON were evaluated using AlmarBlue assay (Invitrogen) over the course of five days, and 48h, respectively, according to the manufacturer’s protocol.

#### Flow cytometry

Activation from latency was measured using GFP (J-Lat 10.6) or HIV-1 core antigen-FITC, KC57 (Beckman Coulter) staining (U1) complemented with Near-IR viability (Invitrogen). Cellular ROS was measured using CellROX™ Deep red Flow Cytometry Assay Kit (Invitrogen), according to the manufacturer’s protocol. Experiments were run on Fortessa flow cytometer (BD Bioscience) and analyzed using FlowJo 10.6.2 (TreeStar Inc).

#### Intracellular metabolite measurement

Intracellular metabolites were measured using Glucose-Glo™ Assay, Lactate-Glo™ Assay, and Glutamate-Glo™ Assay (Promega) according to the manufacturers protocol.

#### Determination of Mitochondrial OXPHOS

U1 and U937 cells (seeding density: 10×10^6^ cells/well) were either left untreated for 48h or treated with DON (12.5µM for 48h), prostratin (untreated for 24h followed by 6µM prostratin for 24h), or DON+Prostratin (12.5µM DON for 48h and 6µM prostratin for last 24h). At 48h, activation of HIV was measured by intracellular staining of HIV-1 core antigen-FITC, KC57 as mentioned earlier. Cells were harvested, washed in PBS, and centrifuged. Cell pellets were processed for mitochondrial extraction using Mitochondria Isolation Kit for Cultured cell (Thermo Scientific) using reagent-based method as per manufacturers guidelines, followed by measuring the OXPHOS suing the total OXPHOS Human WB antibody cocktail (Abcam) with mitochondrial loading control VDAC using the antibody VDAC clone B-6 (Santa Cruz Biotechnology). Relative protein quantification was performed using ImageLab version 6.0.1 (Bio-Rad Laboratories Inc), results analyzed using Mann-Whitney U test or unpaired t-test and visualized using GraphPad Prism 8.4.3 (significance p<0.05). All laboratory experiments were performed in three independent replicates. Analysis was performed using unpaired t-test or Mann-Whitney U test and visualized using GraphPad Prism 8.4.3 (significance *p<0*.*05*).

### Study approval

The Cameroon study was approved by the Cameroon National Ethics Committee for Human Research with Ethical clearance N^O^2019/08-198-CE/CNERSH/SP. The Indian study was approved by the Institutional Ethics Committee of the National Institute for Research in Tuberculosis (NIRT IEC No: 2015023) and the Institutional Review Board of the Government Hospital for Thoracic Medicine (GHTM-27102015) Chennai, India. Ethical approval by Etikprövningsmyndigheten (Sweden) was waived off because of the anonymized data (Dnr 2019-05086). Written informed consent was obtained from all study participants prior inclusion and kept at the respective sites.

## Data Availability

All the data available as supplementary file within manuscript

## Author contributions

Conceptualization and study designing: U.N, Clinical data and biobank: G.M.I., E.L., M.C.O, C.T., H.B. L.E.H, Methodology: F.M., R.B., J.P.M., and U.N., Formal analysis (bioinformatics): M.F. and G.D.V.M. and R.B., Formal analysis (experimental): S.S.A., S.K., M.S., A.E., A.V., and S.G., Supervision: R.B., J.P.M., and U.N., Resources: U.N. Writing (original draft): F.M., S.S.A, S.K., M.S., and UN Writing (review and editing): R.B., G.D.V.M., A.C.B, J.K., K.S., C.L.L., and J.P.M., Visualization: F.M., S.S.A, S.K. and U.N., Project administration: G.M.I., L.E.H. and U.N. Funding acquisition: U.N. All authors discussed the results, commented, and approved the final version of the manuscript.

## Acknowledgments

The study is supported by the Swedish Research Council (2017-01330 and 2018-06156), Karolinska Institutet Stiftelser och Fonder (2020-01554), and Åke Wiberg Stiftelse grant (M18-0021). Swedish Physicians Against AIDS Foundation (FOb20170004) and Jeanssons Stiftelser (JS2016–0185) to UN. MS acknowledges the support received from Swedish Physicians Against AIDS Foundation. We thank all the study subjects for their participation. Authors acknowledge support from the Proteomics Biomedicum; Karolinska Institute, Solna, for LC-MS/MS analysis. Swedish Metabolomics Centre, Umeå, Sweden is acknowledged for targeted metabolic profiling.

## Supplementary Figures

**Figure S1:**
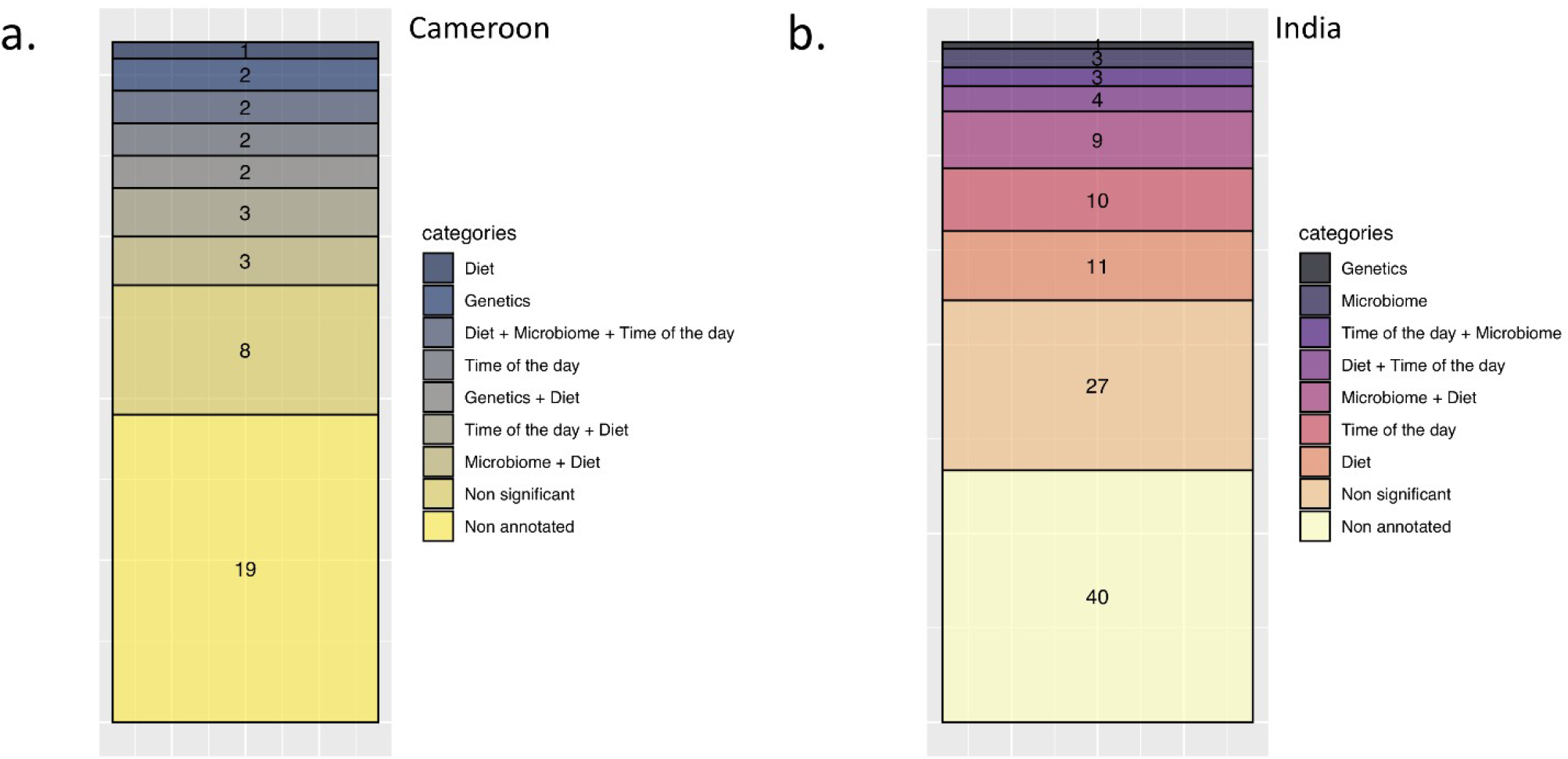
Bar plot representing the number of detected metabolites asociated significantly with each environmental category in Cameroon (a) and India (b) cohorts.

**Figure S2:**
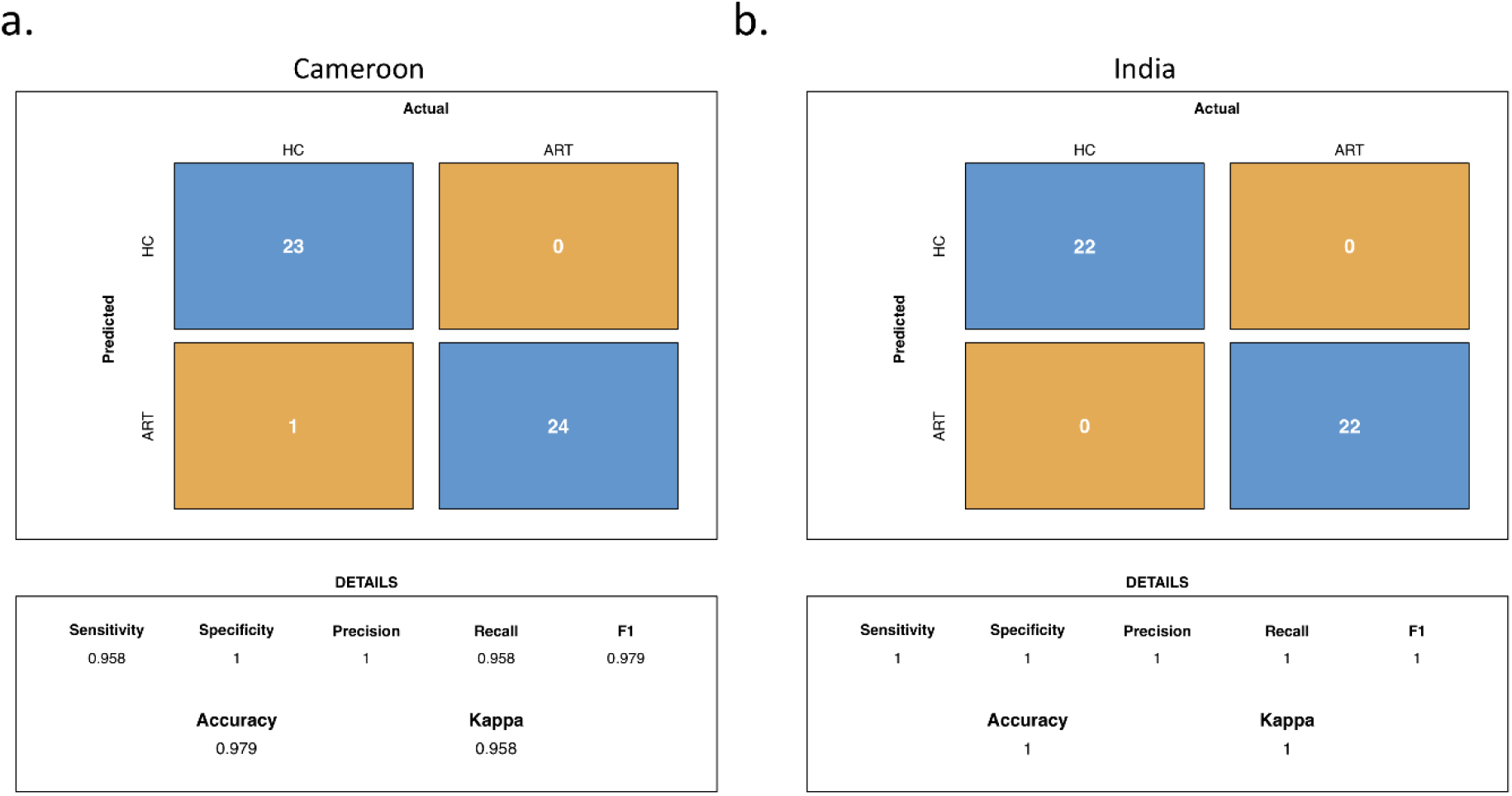
Confusion matrices for Cameroon (A) and India (B) random forest models.

**Figure S3:**
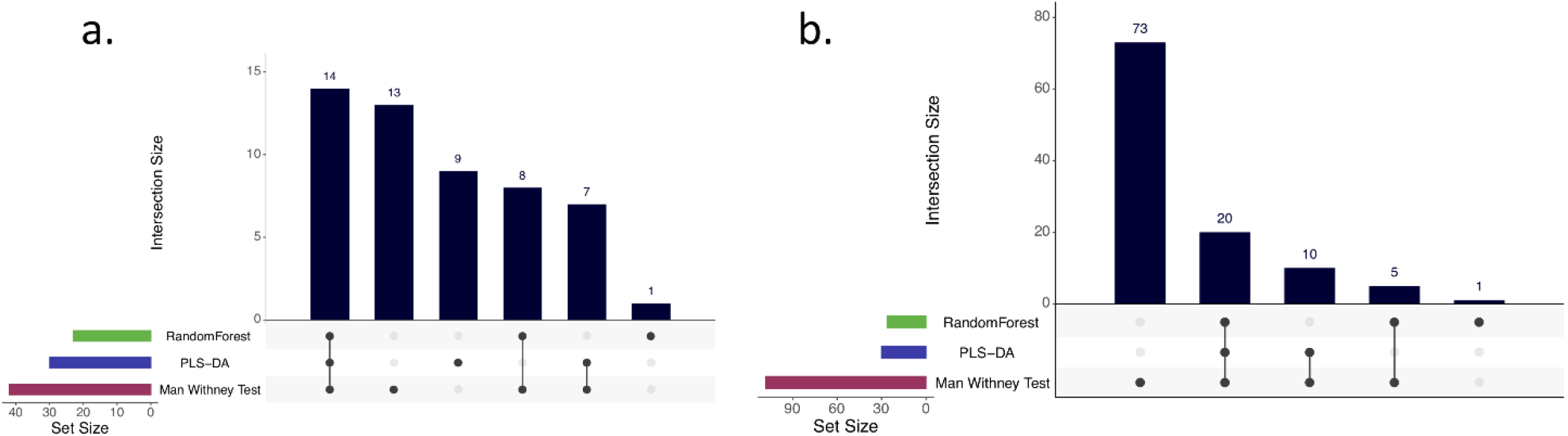
Upset plot of metabolites with differential abundance between HC and ART patients identified by 3 different methods in Cameroon (A) and Indian (B) cohorts. Horizontal bars show the number of metabolites found with each method. Vertical bars display intersects between methods as indicated in the matrix below the graph.

**Figure S4:**
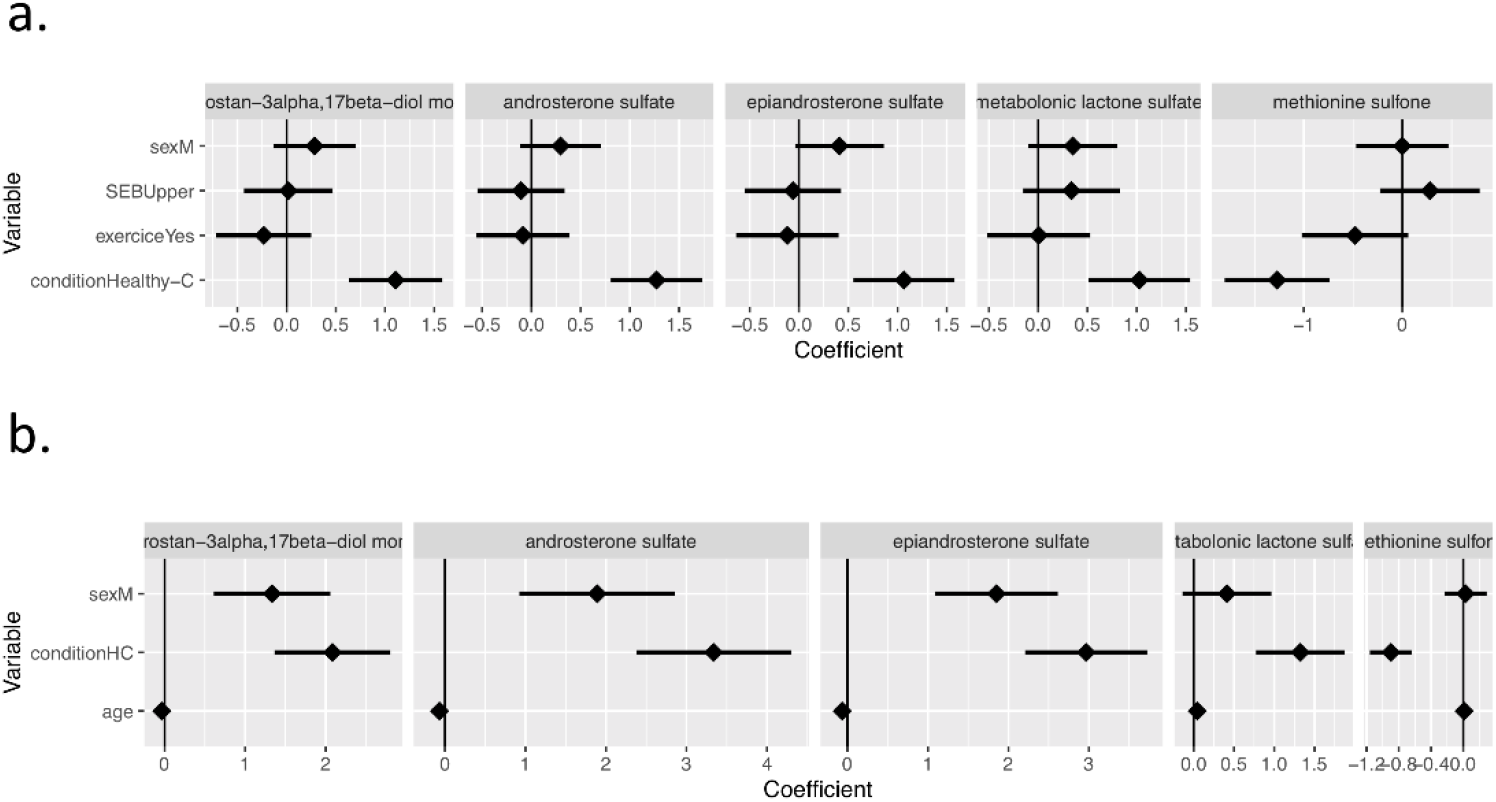
Linear regresion coefficients in Cameroon (a) and Indian (b) cohorts.

**Figure S5:**
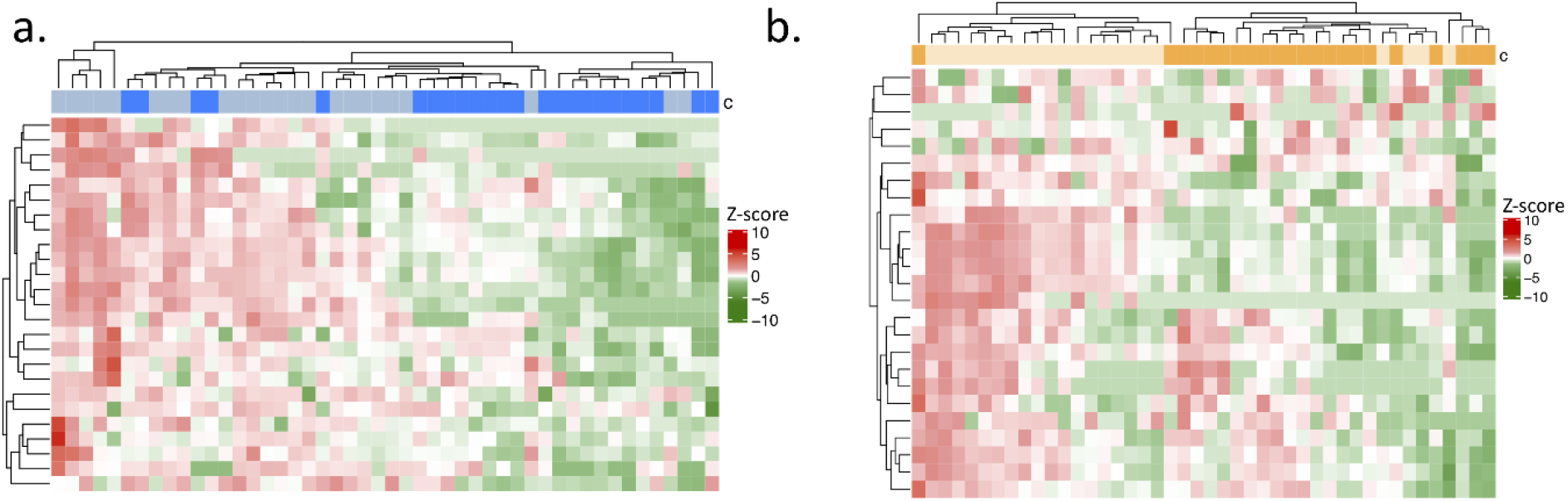
Heatmaps of potential biomarkers and first neighbors in HC and ARTin Cameroon (a) and Indian (b) cohorts.Data were log-transformed and z-score transformed.

**Figure S6:**
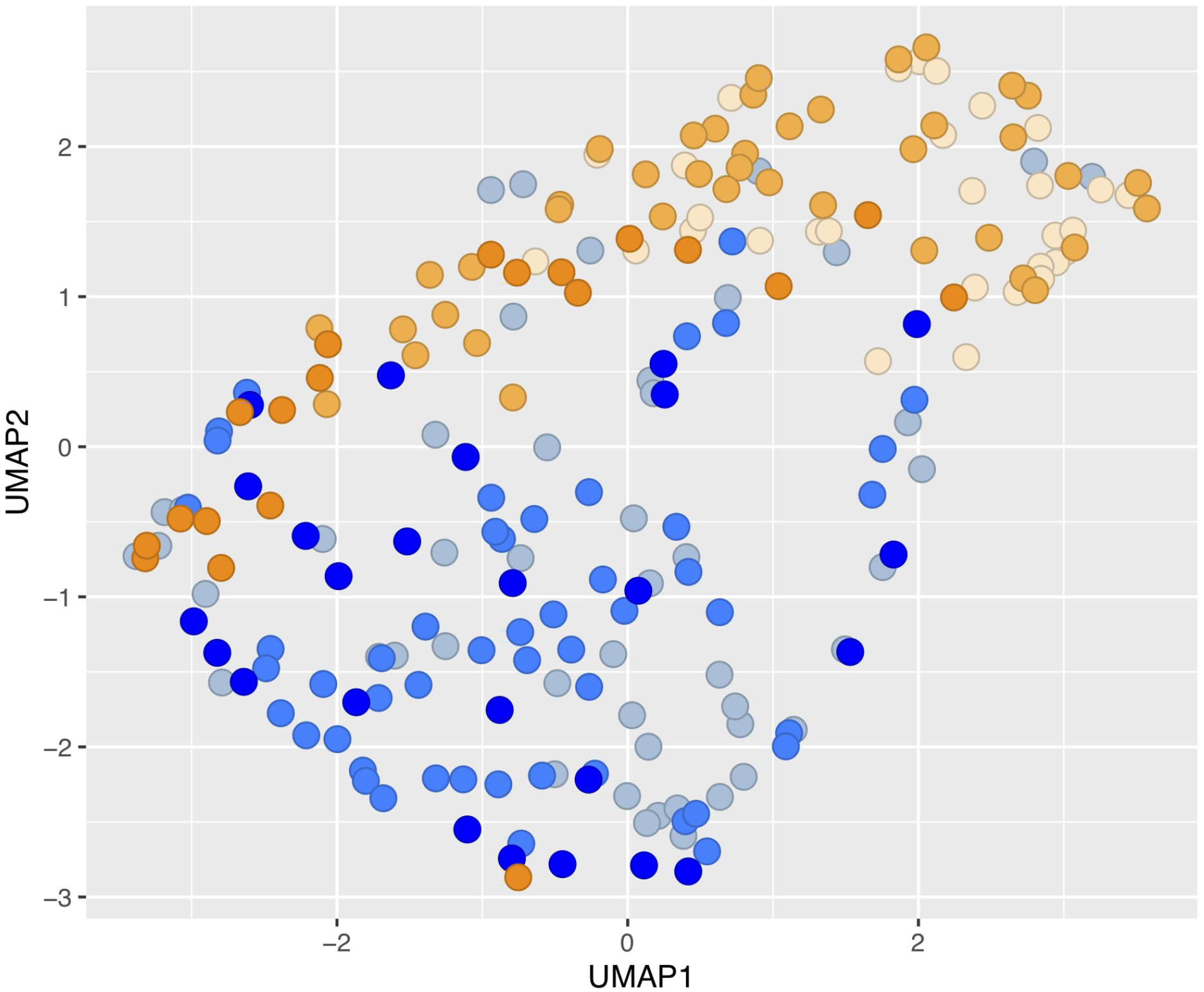
UMAP visualization of AA (targeted metabolomics) for samples from Cameroon and Indian cohorts together.

**Figure S7:**
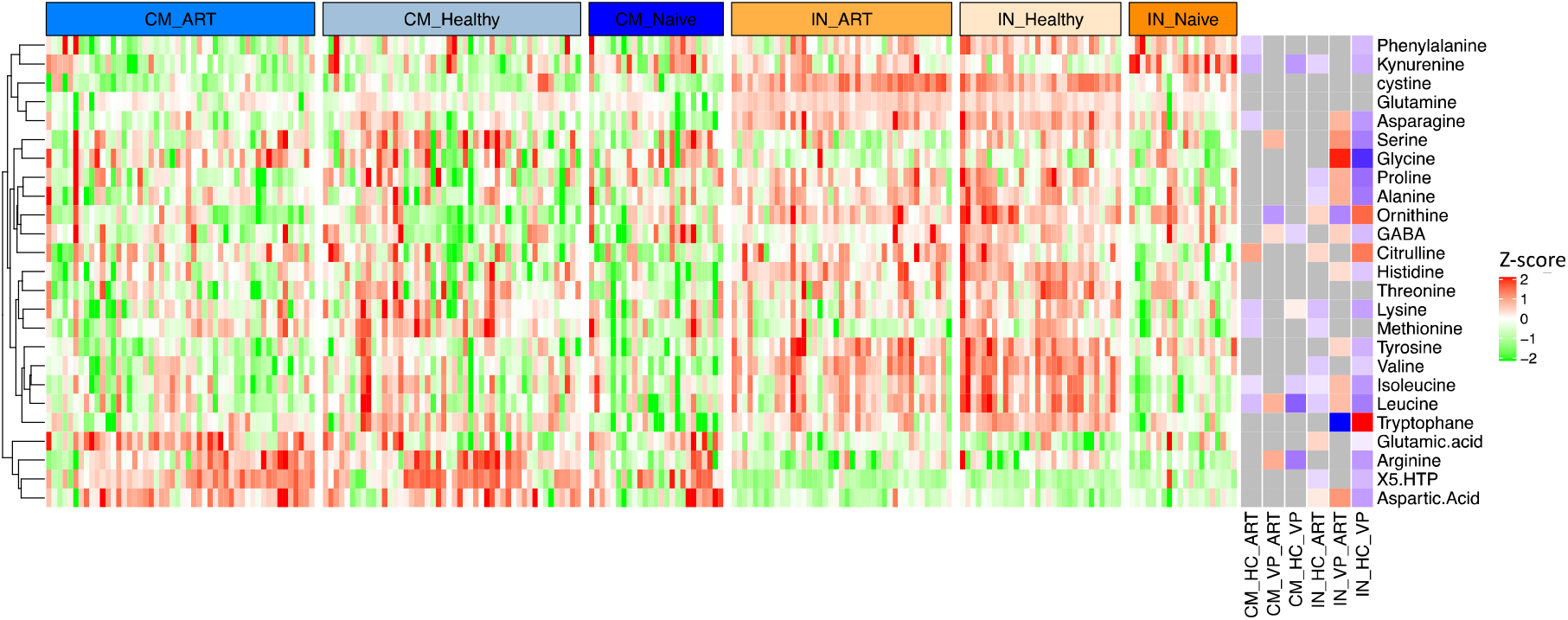
Heatmap targeted metabolomics (aminoacids) for Cameroon and India

**Figure S8:**
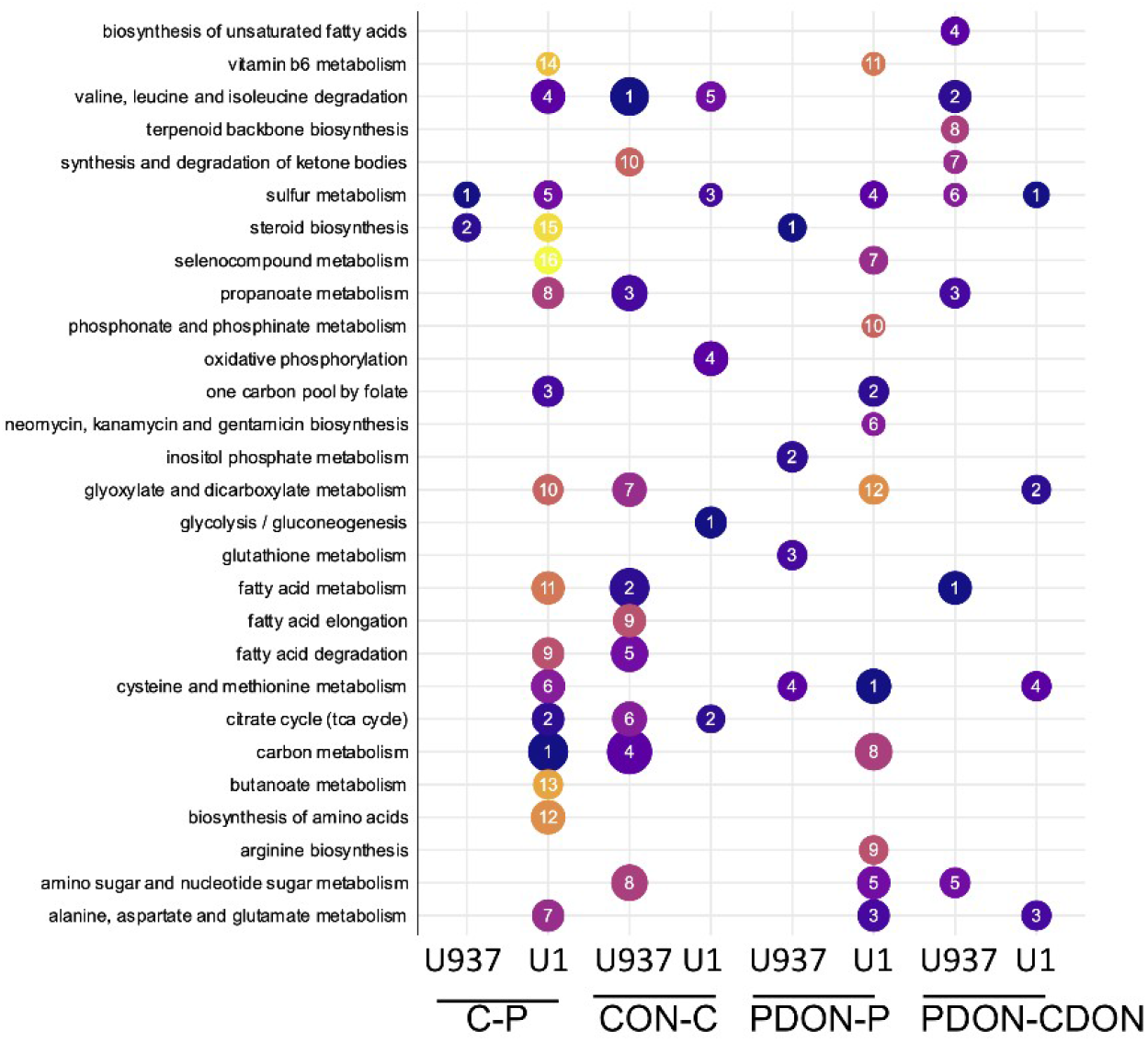
Dot plot representing pathway analysis for all comparison in U937 and U1 corrected for U937 cells. For each cell line, pathways for comparisons Control vs Prostatin (C-P), Control vs DON(CON-C), Prostratin vs Prostratin + DON(PDON-P), DON vs Prostratin + DON(PDON-CDON. Size of the bubble represents the number of proteins and the number,the rank of the pathway based on FDR.

**Figure S9:**
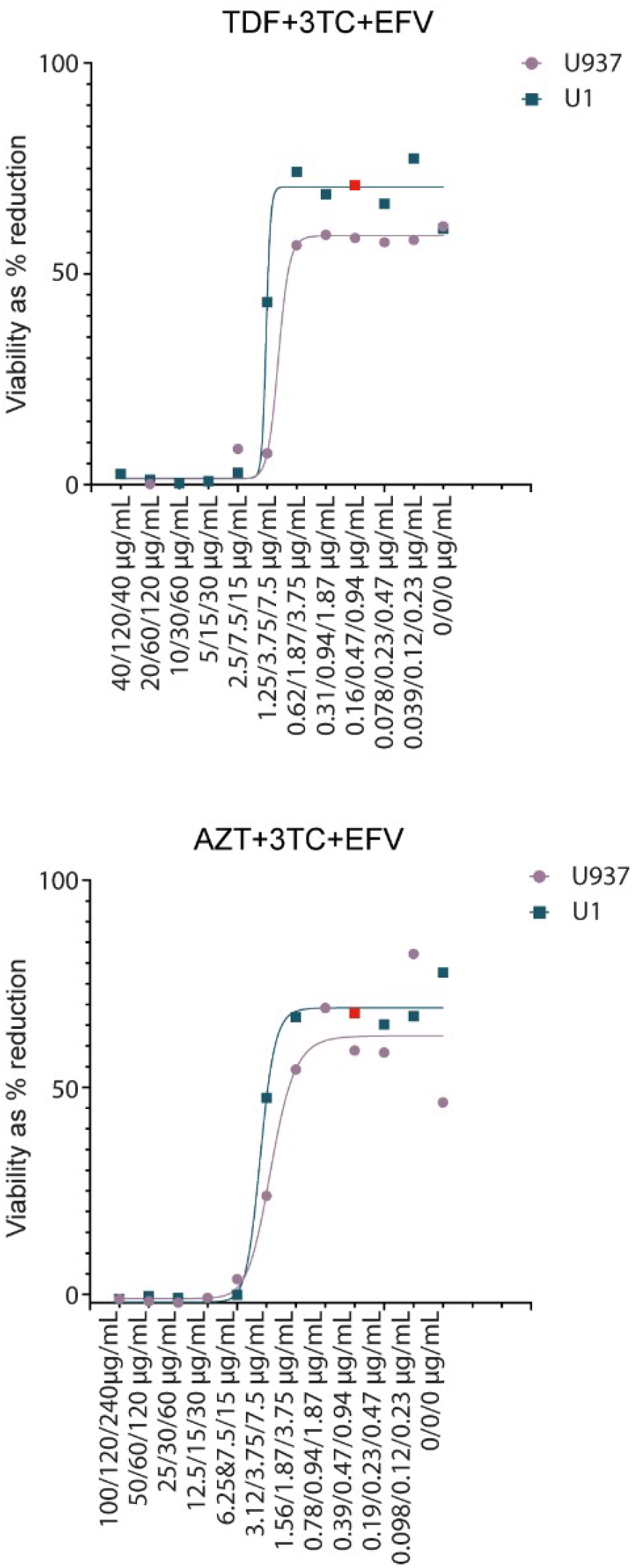
Cytotoxicity of antiretroviral regimens TDF+3TC+EFV and AZT+3TC+EFV in monocytic cell lines,U937 and U1.

**Table S1:** IPA Input list Cameroon.

**Table S2:** Mann Whitney U Test untargeted metabolomics (Cameroon and Indian cohort).

**TableS3:** Mann Whitney U Test targeted metabolomics (Cameroon and Indian cohort).

**TableS4:** Linear regression models adjusted for socio-economic background, exercise and gender showing the association of 6 biomarkers with ART status in the Cameroon cohort.

**TableS5:** Linear regression models adjusted for age and gender showing the association of 6 biomarkers with ART/HC status in the Indian cohort.

**TableS6:** Effect sizes observed between HC and ART (Glass delta).

**TableS7:** Differential protein abundance analysis in U937 and U1 cells using LIMMA.

<u>Supplementary data S1</u>: Normalized untargeted metabolomics data (Cameroon and India) (Metabolon).

<u>Supplementary data S2</u>: Proteomics data after normalization and batch correction (Monocytes).

<u>Supplementary data S3</u>: Proteomics data after normalization and batch correction (Lymphocytes).

## References

1. Babu H, et al. (2019) Plasma Metabolic Signature and Abnormalities in HIV-Infected Individuals on Long-Term Successful Antiretroviral Therapy. Metabolites 9(10).

2. Babu H, et al. (2019) Systemic Inflammation and the Increased Risk of Inflamm-Aging and Age-Associated Diseases in People Living With HIV on Long Term Suppressive Antiretroviral Therapy. Front Immunol 10:1965.

3. Gelpi M, et al. (2021) Central role of the glutamate metabolism in long-term antiretroviral treated HIV-infected individuals with metabolic syndrome: a cross-sectional cohort study. medRxiv:2021.2004.2001.21254778.

4. Rosado-Sánchez I, et al. (2019) Glutaminolysis and lipoproteins are key factors in late immune recovery in successfully treated HIV-infected patients. Clin Sci (Lond) 133(8):997–1010.

5. Cassol E, Misra V, Dutta A, Morgello S, & Gabuzda D (2014) Cerebrospinal fluid metabolomics reveals altered waste clearance and accelerated aging in HIV patients with neurocognitive impairment. Aids 28(11):1579–1591.

6. Munshi SU, Rewari BB, Bhavesh NS, & Jameel S (2013) Nuclear magnetic resonance based profiling of biofluids reveals metabolic dysregulation in HIV-infected persons and those on anti-retroviral therapy. PLoS One 8(5):e64298.

7. Ahmed D, Roy D, & Cassol E (2018) Examining Relationships between Metabolism and Persistent Inflammation in HIV Patients on Antiretroviral Therapy. Mediators Inflamm 2018:6238978.

8. Manchester M & Anand A (2017) Metabolomics: Strategies to Define the Role of Metabolism in Virus Infection and Pathogenesis. Adv Virus Res 98:57–81.

9. Sanchez EL & Lagunoff M (2015) Viral activation of cellular metabolism. Virology 479-480:609–618.

10. Sigal A, et al. (2011) Cell-to-cell spread of HIV permits ongoing replication despite antiretroviral therapy. Nature 477(7362):95–98.

11. Bar N, et al. (2020) A reference map of potential determinants for the human serum metabolome. Nature 588(7836):135–140.

12. Becker S, Kortz L, Helmschrodt C, Thiery J, & Ceglarek U (2012) LC-MS-based metabolomics in the clinical laboratory. J Chromatogr B Analyt Technol Biomed Life Sci 883-884:68–75.

13. Yet I, et al. (2016) Genetic Influences on Metabolite Levels: A Comparison across Metabolomic Platforms. PLoS One 11(4):e0153672.

14. Valle-Casuso JC, et al. (2019) Cellular metabolism is a major determinant of HIV-1 reservoir seeding in CD4+ T cells and offers an opportunity to tackle infection. Cell metabolism 29(3):611-626. e615.

15. Clerc I, et al. (2019) Entry of glucose-and glutamine-derived carbons into the citric acid cycle supports early steps of HIV-1 infection in CD4 T cells. Nature metabolism 1(7):717–730.

16. Cassol E, et al. (2013) Plasma metabolomics identifies lipid abnormalities linked to markers of inflammation, microbial translocation, and hepatic function in HIV patients receiving protease inhibitors. BMC Infect Dis 13:203.

17. Peltenburg NC, et al. (2018) Persistent metabolic changes in HIV-infected patients during the first year of combination antiretroviral therapy. Sci Rep 8(1):16947.

18. Ziegler TR, Judd SE, Ruff JH, McComsey GA, & Eckard AR (2017) Amino Acid Concentrations in HIV-Infected Youth Compared to Healthy Controls and Associations with CD4 Counts and Inflammation. AIDS Res Hum Retroviruses 33(7):681–689.

19. Sitole LJ, Tugizimana F, & Meyer D (2019) Multi-platform metabonomics unravel amino acids as markers of HIV/combination antiretroviral therapy-induced oxidative stress. J Pharm Biomed Anal 176:112796.

20. Williams AA, Sitole LJ, & Meyer D (2017) HIV/HAART-associated oxidative stress is detectable by metabonomics. Mol Biosyst 13(11):2202–2217.

21. Binvignat O & Olloquequi J (2020) Excitotoxicity as a Target Against Neurodegenerative Processes. Curr Pharm Des 26(12):1251–1262.

22. Eck HP, Frey H, & Dröge W (1989) Elevated plasma glutamate concentrations in HIV-1-infected patients may contribute to loss of macrophage and lymphocyte functions. Int Immunol 1(4):367–372.

23. Gorska AM & Eugenin EA (2020) The Glutamate System as a Crucial Regulator of CNS Toxicity and Survival of HIV Reservoirs. Front Cell Infect Microbiol 10:261.

24. Hegedus A, et al. (2017) Evidence for Altered Glutamine Metabolism in Human Immunodeficiency Virus Type 1 Infected Primary Human CD4(+) T Cells. AIDS Res Hum Retroviruses 33(12):1236–1247.

25. Zhao J, et al. (2004) Mitochondrial glutaminase enhances extracellular glutamate production in HIV-1-infected macrophages: linkage to HIV-1 associated dementia. J Neurochem 88(1):169–180.

26. Nedelcovych MT, et al. (2019) Glutamine Antagonist JHU083 Normalizes Aberrant Glutamate Production and Cognitive Deficits in the EcoHIV Murine Model of HIV-Associated Neurocognitive Disorders. J Neuroimmune Pharmacol 14(3):391–400.

27. Nedelcovych MT, et al. (2017) N-(Pivaloyloxy)alkoxy-carbonyl Prodrugs of the Glutamine Antagonist 6-Diazo-5-oxo-l-norleucine (DON) as a Potential Treatment for HIV Associated Neurocognitive Disorders. J Med Chem 60(16):7186–7198.

28. Mukerji SS, et al. (2021) Low Neuroactive Steroids Identifies a Biological Subtype of Depression in Adults with Human Immunodeficiency Virus on Suppressive Antiretroviral Therapy. J Infect Dis 223(9):1601–1611.

29. Giatti S, Garcia-Segura LM, Barreto GE, & Melcangi RC (2019) Neuroactive steroids, neurosteroidogenesis and sex. Prog Neurobiol 176:1–17.

30. Valle-Casuso JC, et al. (2019) Cellular Metabolism Is a Major Determinant of HIV-1 Reservoir Seeding in CD4(+) T Cells and Offers an Opportunity to Tackle Infection. Cell Metab 29(3):611–626 e615.

31. Clerc I, et al. (2019) Entry of glucose- and glutamine-derived carbons into the citric acid cycle supports early steps of HIV-1 infection in CD4 T cells. Nature Metabolism 1(7):717–730.

32. Hegedus A, et al. (2017) Evidence for Altered Glutamine Metabolism in Human Immunodeficiency Virus Type 1 Infected Primary Human CD4(+) T Cells. AIDS research and human retroviruses 33(12):1236–1247.

33. Datta PK, et al. (2016) Glutamate metabolism in HIV-1 infected macrophages: Role of HIV-1 Vpr. Cell cycle (Georgetown, Tex.) 15(17):2288–2298.

34. Huang Y, et al. (2011) Glutaminase Dysregulation in HIV-1-Infected Human Microglia Mediates Neurotoxicity: Relevant to HIV-1-Associated Neurocognitive Disorders. The Journal of Neuroscience 31(42):15195.

35. Castellano P, Prevedel L, Valdebenito S, & Eugenin EA (2019) HIV infection and latency induce a unique metabolic signature in human macrophages. Sci Rep 9(1):3941.

36. Loisel-Meyer S, et al. (2012) Glut1-mediated glucose transport regulates HIV infection. Proceedings of the National Academy of Sciences of the United States of America 109(7):2549–2554.

37. Palmer CS, et al. (2017) Metabolically active CD4+ T cells expressing Glut1 and OX40 preferentially harbor HIV during in vitro infection. FEBS Lett 591(20):3319–3332.

38. Palmer CS, et al. (2014) Increased glucose metabolic activity is associated with CD4+ T-cell activation and depletion during chronic HIV infection. AIDS 28(3):297–309.

39. Palmer CS, et al. (2014) Glucose Transporter 1–Expressing Proinflammatory Monocytes Are Elevated in Combination Antiretroviral Therapy–Treated and Untreated HIV+ Subjects. The Journal of Immunology 193(11):5595–5603.

40. Sperk M, et al. (2021) Distinct lipid profile, low-level inflammation, and increased antioxidant defense signature in HIV-1 elite control status. iScience 24(2):102111.

41. Appelberg S, et al. (2020) Dysregulation in Akt/mTOR/HIF-1 signaling identified by proteo-transcriptomics of SARS-CoV-2 infected cells. Emerg Microbes Infect 9(1):1748–1760.

42. Kursa MB & Rudnicki WR (2010) Feature selection with the Boruta package: Journal.

43. Pedregosa F, et al. (2011) Scikit-learn: Machine learning in Python. the Journal of machine Learning research 12:2825–2830.

44. Thévenot EA, Roux A, Xu Y, Ezan E, & Junot C (2015) Analysis of the Human Adult Urinary Metabolome Variations with Age, Body Mass Index, and Gender by Implementing a Comprehensive Workflow for Univariate and OPLS Statistical Analyses. J Proteome Res 14(8):3322–3335.

45. Ritchie ME, et al. (2015) limma powers differential expression analyses for RNA-sequencing and microarray studies. Nucleic Acids Res 43(7):e47.

46. Traag VA, Waltman L, & van Eck NJ (2019) From Louvain to Leiden: guaranteeing well-connected communities. Sci Rep 9(1):5233.

47. Wilkerson MD & Hayes DN (2010) ConsensusClusterPlus: a class discovery tool with confidence assessments and item tracking. Bioinformatics 26(12):1572–1573.

48. Leland M, John H, Nathaniel S, & Lukas G (2018) UMAP: uniform manifold approximation and projection. Journal of Open Source Software 3(29):861.

49. Wickham H (2016) ggplot2. Elegant Graphics for Data Analysis (Springer International Publishing, New York) pp XVI, 260.

50. Brunson JC (2020) Ggalluvial: layered grammar for alluvial plots. Journal of Open Source Software 5(49):2017.

51. Pérez-Silva JG, Araujo-Voces M, & Quesada V (2018) nVenn: generalized, quasi-proportional Venn and Euler diagrams. Bioinformatics 34(13):2322–2324.

52. Chen H & Boutros PC (2011) VennDiagram: a package for the generation of highly-customizable Venn and Euler diagrams in R. BMC bioinformatics 12(1):1–7.

53. Conway JR, Lex A, & Gehlenborg N (2017) UpSetR: an R package for the visualization of intersecting sets and their properties. Bioinformatics 33(18):2938–2940.

54. Gu Z, Eils R, & Schlesner M (2016) Complex heatmaps reveal patterns and correlations in multidimensional genomic data. Bioinformatics 32(18):2847–2849.

55. Shannon P, et al. (2003) Cytoscape: a software environment for integrated models of biomolecular interaction networks. Genome Res 13(11):2498–2504.

